# Molecular diagnosis of causality in T cell mediated severe cutaneous adverse drug reactions

**DOI:** 10.1101/2021.02.16.21250166

**Authors:** Ying X Teo, Wei Y Haw, Andres F Vallejo, Carolann McGuire, Jeongmin Woo, Peter S Friedmann, Marta E Polak, Michael R Ardern-Jones

## Abstract

**Background:** One of the most severe forms of T cell mediated cutaneous adverse drug reactions is ‘drug reaction with eosinophilia and systemic symptoms’ (DRESS), hence subsequent avoidance of the causal drug is imperative. However, attribution of drug culpability in DRESS is challenging and standard skin allergy tests are not recommended due to for patient safety reasons. We sought to identify potential biomarkers for development of a diagnostic test.

**Methods:** Peripheral blood mononuclear cells (PBMCs) from a ‘discovery’ cohort (n=5) challenged to drug or control were analysed for transcriptomic profile. A signature panel of genes was then tested in a validation cohort (n=6), and compared to tolerant controls and other inflammatory conditions which can clinically mimic DRESS. A scoring system to identify presence of drug hypersensitivity was developed based on gene expression alterations of this panel.

**Results:** Whole transcriptome analysis identified 4 major gene clusters including those regulating T cell activation via NFAT and cytokine receptor activity. 22 differentially expressed gene transcripts were identified as a DRESS signature including Type 1 interferon pathways and Th2 activation. The DRESS transcriptomic panel identified antibiotic-DRESS cases in a validation cohort but was not altered in other inflammatory conditions. Machine learning or differential expression selection of a biomarker panel showed high sensitivity and specificity (100% and 85.7-100% respectively) for identification of the culprit drug in DRESS.

**Conclusion:** Transcriptomic analysis of DRESS revealed important insights into the key activated pathways and identified a transcriptional signature which shows potential as a test with high sensitivity for drug culpability attribution.

## Introduction

Drug allergies (drug hypersensitivity) caused by T cell mediated reactions are clinically distinct in their presentation from IgE-mediated drug allergy reactions and present as a range of different clinical phenotypes (1), including Drug Reaction with Eosinophilia and Systemic Symptoms (DRESS). DRESS typically presents with a florid skin eruption combined with hallmark systemic features of fever, lymphadenopathy, blood dyscrasias such as eosinophilia, and internal organ involvement (1–3). The liver is the most commonly involved among the organs, found in 51–94.2% of patients; followed by renal involvement, lung, cardiac and central nervous system (4–7). Future lifelong avoidance of the culprit drug is crucial as DRESS can be life-threatening, reported mortality being 2-6% (2, 4, 8). Confirmation of causality can be difficult if the culprit drug is not clinically obvious.

Skin tests and oral challenge cannot be performed acutely and are generally not recommended because of the risk of re-inducing DRESS. Clinical algorithms to assess causality are of value, especially for post-marketing surveillance systems, but their lack of confirmatory testing limits their utility to inform treatment decisions for an individual patient (9). We and others have demonstrated the diagnostic use of classical immunology tests to measure drug specific T cell activation (10, 11). However, such *in vitro* assays are not widely available due to being labour intensive, complex, and involving radioisotopes. Therefore, there is an unmet need to develop a simple, quick and robust *in vitro* assay that can be undertaken widely in routine diagnostic laboratories.

We set out to develop an *in vitro* gene transcription signature to identify drug-induced T cell activation because reverse transcriptase polymerase chain reaction (RT-PCR) based assays are already widely employed in clinical laboratories and therefore this approach would be scalable to routine laboratories. To determine the optimal biomarkers for drug T cell activation, we undertook ribonucleic acid-sequencing (RNA-seq) of drug-exposed peripheral blood mononuclear cells (PBMCs) from antibiotic-induced DRESS cases. Differential expression from control samples identified candidate genes as markers of drug hypersensitivity, which were further validated against a second cohort, against tolerant controls and in other inflammatory conditions.

## Results

### Correlation between clinical diagnosis and *in vitro* assays

DRESS was the most common presentation of DHR in Southampton tertiary referral centre (53% of diagnosed DHR in 2017-2018) and in our cohort, antibiotics were the dominant causal drugs for this condition (Figure 1a). 5 cases of antibiotic-induced DRESS were selected (‘discovery’ cohort). Cohort characteristics (median age 49 years, IQR: 36-71), are described in Table 1. We confirmed that all identified antibiotic-DRESS cases demonstrated positive *in vitro* responses to stimulation with the culprit antibiotic, whereas no drug-induced responses were detected in tolerant controls (LPA p = 0.0025, IFN-γ p =0.0025, Mann-Whitney U test) (Figure 1 b,c).

**Table 1:**
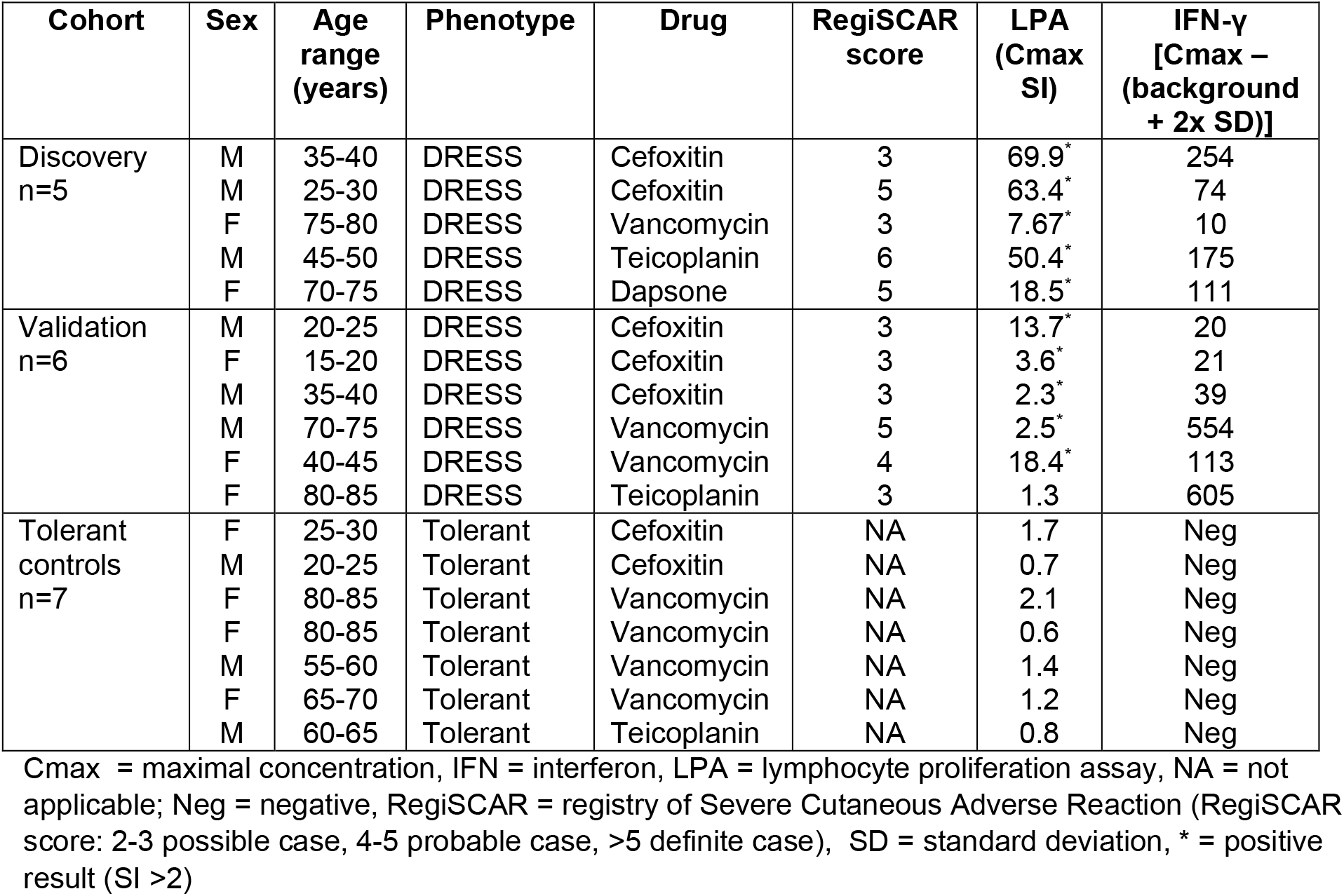
Demographics of tested subjects and comparative T-cell assay results

**Figure 1:**
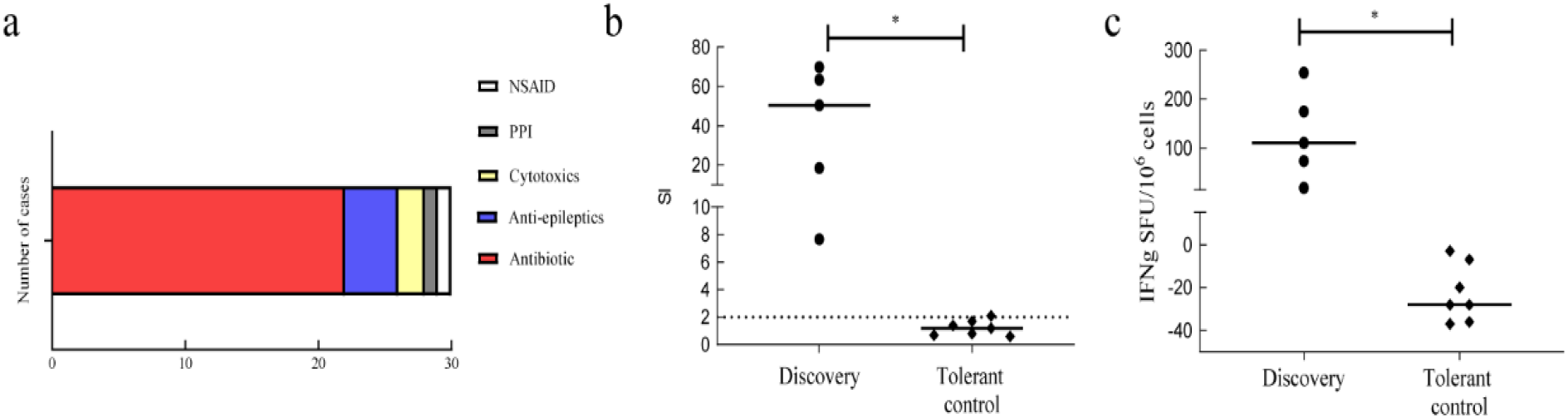
Causative drugs in referred DRESS cases and confirmation of clinically suspected antibiotic by positive T cell assay in DRESS ‘discovery’ cohort. a) Prevalence of causative drug groups in DRESS cases referred to Southampton NHS Foundation Trust between 2017-2018. b) Lymphocyte Proliferation Test (LPA) measured as stimulation index (SI) of proliferation induced by drug versus media control and c) IFN-γ release in drug-induced responses measured by ELIspot in ‘discovery’ cohort subjects (n=5), and control patients tolerant of similar antibiotics (n=7). Each data point represents maximum measured response to tested drug. Horizontal solid lines indicate group median. Horizontal dotted line shows positive result threshold. Mann-Whitney U test used for assessing statistical significance, * = p-value <0.05. DRESS = drug reaction with eosinophilia and systemic symptoms, IFN-γ ELIspot = interferon-gamma enzyme-linked immunosorbent spot, LPA = lymphocyte proliferation assay, NSAID = non-steroidal anti-inflammatory, PPI = protein pump inhibitor, SFU = spot forming unit

### Antibiotic exposure induces transcriptomic programmes encoding immune activation in PBMCs from DRESS patients

To identify transcriptomic biomarkers specific for DRESS induced by antibiotics, discovery cohort PBMCs were co-cultured with culprit drug or control *in vitro* for 24 hours before isolation of RNA for transcriptome profiling (Figure 2a). This identified 267 drug-specific differentially expressed genes (DEGs) (149 up and 118 down-regulated; EdgeR, FDR p<0.05, logFC ≥|1|, Figure 2b). Transcript-to-transcript clustering (GraphiaPro, Pearson r≥0.85, MCL = 1.7) identified 4 main clusters (Figure 2c). Clusters 1 and 3, comprising 141 genes in total, were enriched in genes regulating cytokine receptor activity (Cluster 1, FDR p =7.67×10^−7^) and T cell activation via NFAT (Cluster 3, FDR p =1×10^−3^, Figure 2d,e). In contrast, genes in clusters 2 and 4 were downregulated, and indicated modulation of innate immune system function (Cluster 2, FDR p =1.87×10^−2^) and reduced integrin interactions (Cluster 4, FDR p = 1.65 ×10^−3^, Figure 2d,e).

**Figure 2:**
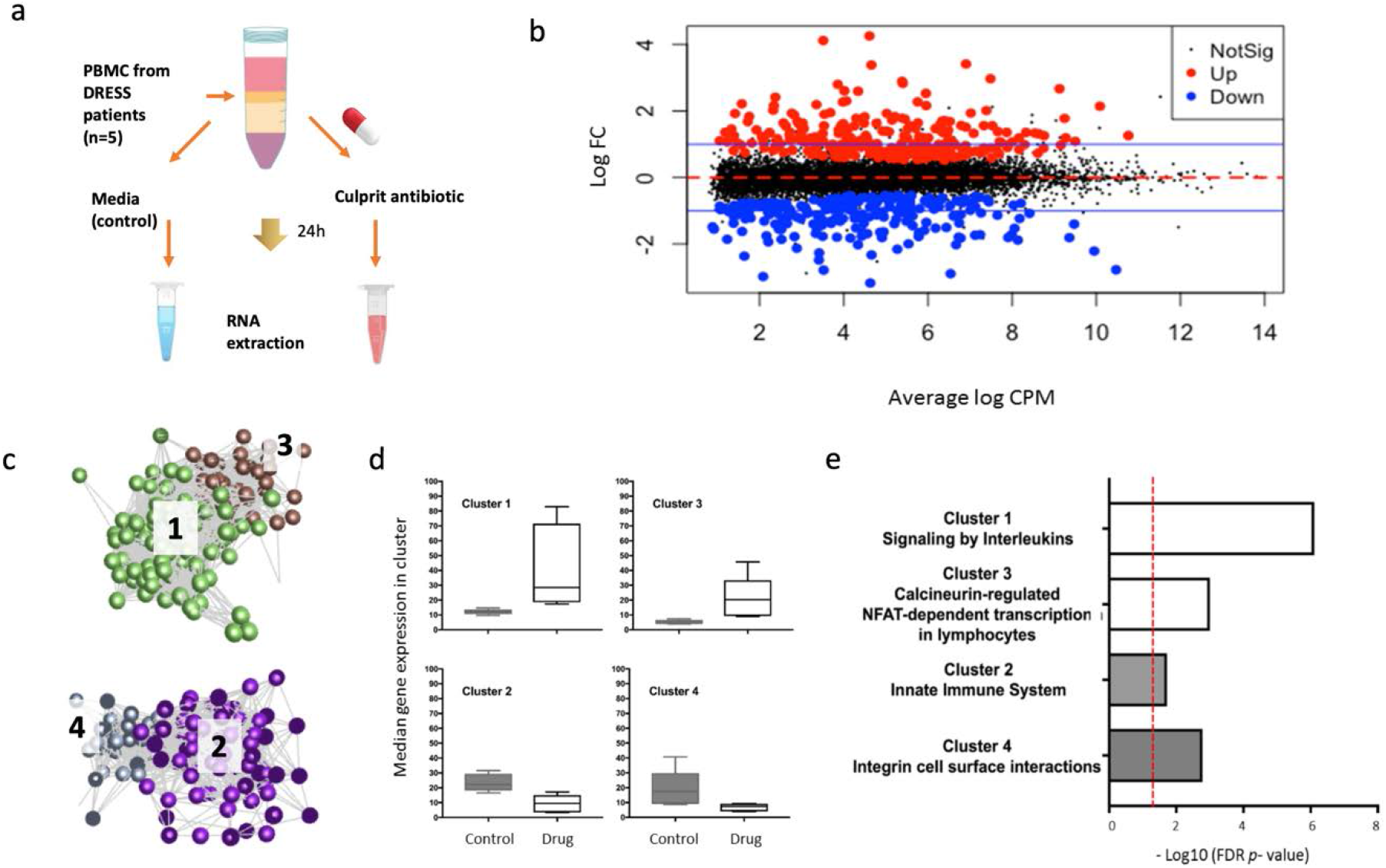
Testing protocol and identification of Differentially Expressed Genes. a) PBMCs were cultured in culture media supplemented or not with culprit drug at the optimised concentration for 24 hours before RNA extraction. b) MA plot representation of 267 drug-specific DEGs (149 up-regulated, red; 118 down-regulated, blue; FDR p<0.05, blue line depicts a threshold of logFC ≥|1|). c-e) Transcript-to-transcript correlation network analysis of gene expression changes induced by culprit drug in DRESS patients (discovery cohort, n=5). 4 major clusters shown, cluster 1 (green, n=103 genes), cluster 2 (purple, n= 76 genes), cluster 3 (brown, n=39 genes), cluster 4 (grey, n = 32 genes). Each node (dot) indicates a transcript, each line defines the Pearson correlation coefficient between a pair of nodes (GraphiaPro, Pearson r≥0.85, MCL = 1.7). d) Median gene expression profiles in clusters 1-4 in control (grey) and drug exposed cells (white). Box and whiskers indicate median +/-range. e) Key processes identified by gene ontology analysis specific to each cluster (ToppGene, FDR cut-off 0.05, cluster 1: FDR p =7.67×10^−7^, cluster 2: FDR p=1.87×10^−2^; cluster 3: FDR p=1×10^−3^; cluster 4: FDR p = 1.65 ×10^−3^). DEG=differentially expressed gene, FC=fold change, FDR=false discovery rate

### Identification of candidate molecular biomarkers for DRESS

To select a panel of candidate biomarkers, DEGs exceeding |logFC| ≥ 1.5 were filtered for the nominal gene expression value (minimum cpm ≥ 4 for all the donors, at least 100cpm). The resulting 48 candidate biomarkers were evaluated for predictive value using a random forest algorithm in R (package Ranger, alpha=0.9, trees = 500). The top 10 genes with absolute FC ≥ |2| (up and down regulation) and RF importance ≥ 0.05 (Figure 3a, b) and 12 additional immune-related genes were included in the final candidate biomarker panel (Figure 3a, full list of genes including 2 housekeeping genes in Supplemental Table S1). Unsupervised principal component clustering of the candidate biomarkers confirmed that they efficiently differentiated drug-exposed cells from their media control counterparts (Figure 3c). RNA-seq analysis was validated using RT-qPCR for the top 4 gene transcripts (Supplemental Figure 1) and a customised array card confirming the differential expression profile of all 22 transcripts ((r = 0.9542 p = <0.0001) Figure 3d). The differential expression of the candidate biomarker panel (Figure 3e) highlights that although the signature differentiates drug-exposed cells from the control, a degree of heterogeneity existed in expression of specific genes between different subjects.

**Figure 3:**
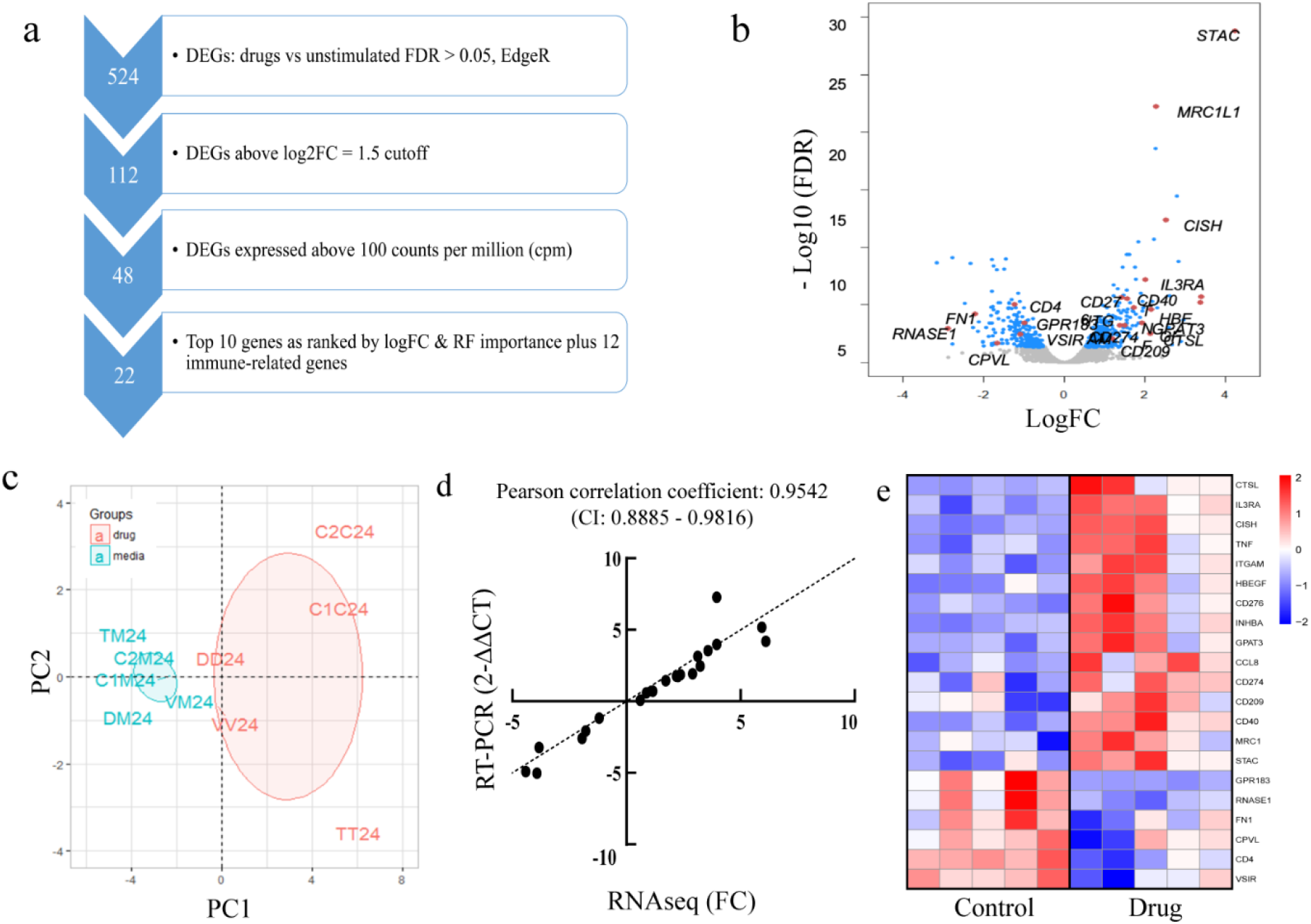
Identification of candidate biomarker genes. a) Selection of candidate biomarkers. Following identification of 524 DEGs by comparison of drug-stimulated and media (unstimulated) in the discovery cohort (EdgeR package, FDR<0.05), genes with |logFC| ≥ 1.5 and cpm >100 were selected. 10 genes with absolute FC ≥ |2|and random forest (RF) importance ≥ 0.05 were selected from the filtered genes and combined with 12 immune-related genes to form the gene panel. b) Volcano plot of genes measured in DRESS discovery cohort, differentiating responses to culprit drug versus media control. Differentially expressed genes (FDR<0.05) shown in blue (up-regulated genes on right, down-regulated on left), genes selected indicated in red. c) PCA clustering (first two components) comparing signature panel gene expression induced by culprit drug (red) and media (blue) after 24-hour culture. d) Comparison of gene changes detected in panel genes using RNAseq and PCR with customised microfluidic array card in a single subject, normalised to YWHAZ gene expression. e) Heatmap depicting changes in expression of selected 22 candidate biomarkers in 5 antibiotic-DRESS patients exposed to culprit drug versus media control. Colour indicates the expression change in logFC. Red: upregulated genes; blue: downregulated genes. DEG=differentially expressed gene, FC=fold change, PC = principal component, PCA = principal component analysis, RF = random forest

### DRESS biomarkers are specific to drug hypersensitivity

To determine if the identified biomarker panel was DRESS specific, we undertook a comparative analysis with influenza infection (GSE114588), sepsis (GSE60424), systemic lupus erythematosus (GSE112087) and dermatomyositis (GSE125977). Gene expression in these four conditions differed markedly from DRESS (Figure 4), and showed low correlations between DRESS and influenza (0.351), sepsis (−0.179), systemic lupus erythematosus (0.327), and dermatomyositis (0.321) (Pearson correlation coefficient).

**Figure 4.**
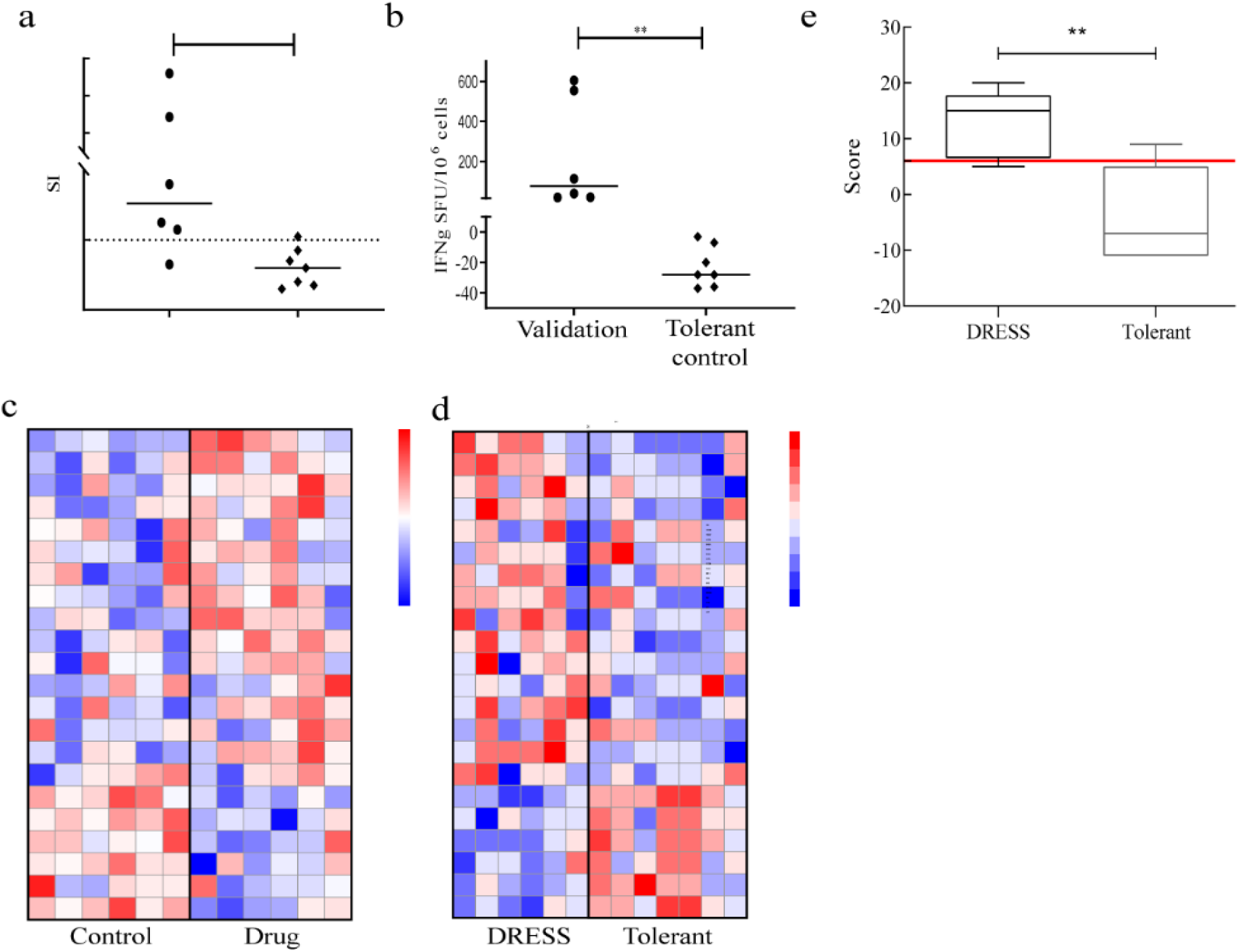
DRESS biomarkers are specific to drug hypersensitivity. Heatmap depicting expression of biomarker gene panel in samples sourced from public data repositories including influenza infection (GSE114588), sepsis (GSE60424), systemic lupus erythematosus (GSE112087) and dermatomyositis (GSE125977). Colour indicates the expression change compared to DRESS allergics. Red: upregulated genes, Blue: downregulated genes.

### Validation of DRESS gene signature panel

To confirm the candidate molecular biomarker panel, we prospectively identified a ‘validation cohort’ (6 cases of DRESS caused by antibiotics: cefoxitin, vancomycin and teicoplanin) as well as patients tolerant of the same antibiotics (n=7). This group was similar in terms of age, sex, and time to onset (Table 1). Similar to the discovery cohort, positive tests for drug hypersensitivity were demonstrated by T cell functional assays *in vitro* (LPA p = 0.0082, IFN-γ p =0.0012, Mann-Whitney U test) in all DRESS subjects (Figure 5a, b). To validate the gene signature panel, PBMCs from allergics were challenged with culprit medications, and the 22-candidate biomarker panel analysed. Comparison of culprit drug against media control in DRESS patients (Figure 5c) and between DRESS cohort against tolerant individuals (Figure 5d) showed clearly identifiable differences. In tolerant subjects, the 22 candidate biomarkers tested were only minimally affected following exposure to antibiotics (median change in gene expression relative to *YWHAZ* for each gene 2^−ΔΔCT^ = 1.04, range: 0.68-1.81), confirming the signature was specific for DRESS. As expected, some heterogeneity in the gene expression patterns between individuals was evident in both tolerant controls and allergic individuals.

**Figure 5.**
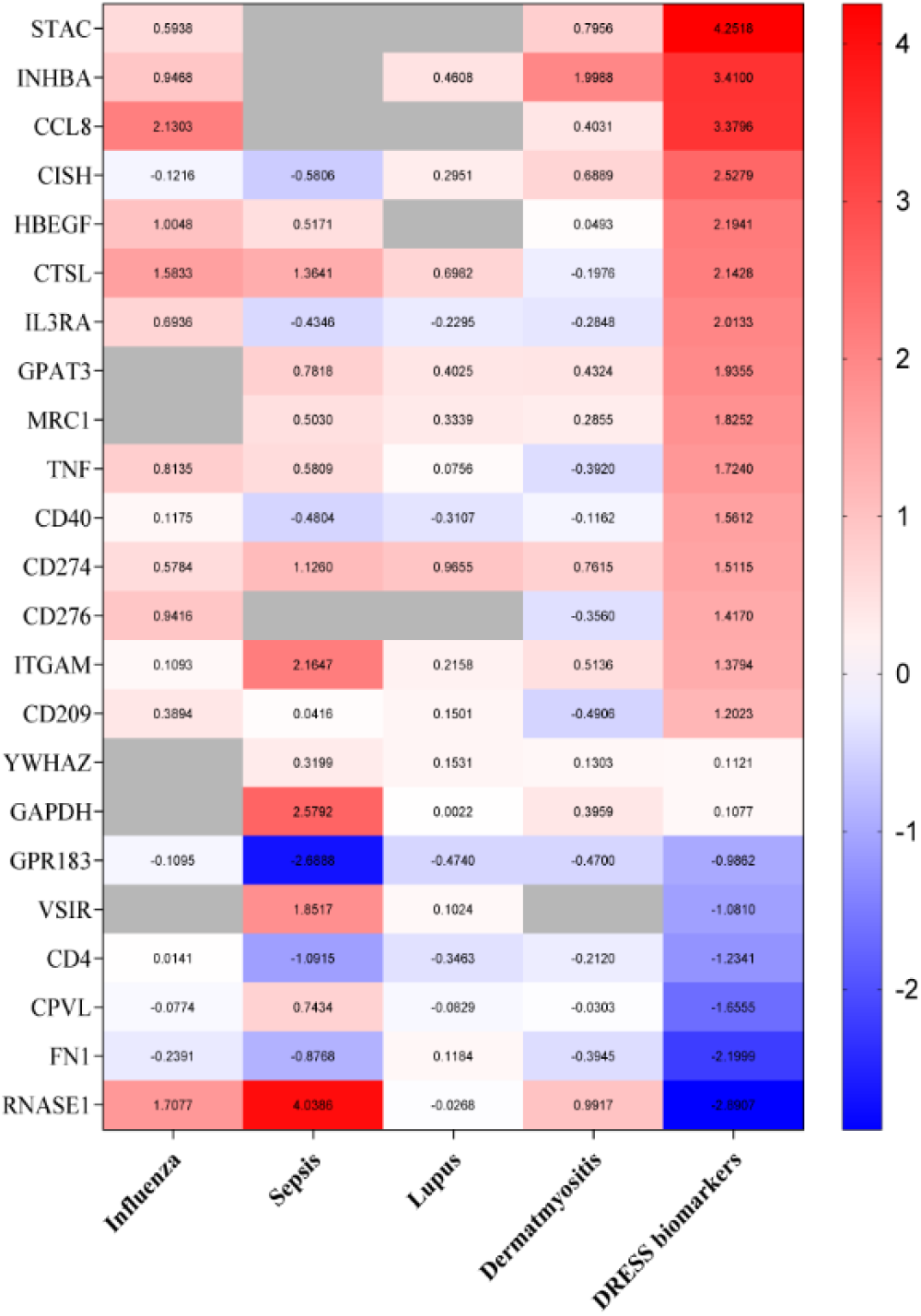
DRESS validation cohort confirms specificity of biomarker panel. a-b) Characteristics of *in vitro* responses to culprit drug in antibiotic-DRESS validation cohort (n=6) and control patients tolerant of similar antibiotics (n=7). a) Lymphocyte Proliferation Test (LPA) measured as stimulation index (SI) of proliferation induced by drug versus media control and b) IFN-γ release in drug-induced responses measured by ELISpot. Each data point represents maximum measured response to tested drug. Each data point represents maximum measured response to tested drug. Horizontal solid lines indicate group median. Horizontal dotted line shows positive result threshold. Mann-Whitney U test used for statistical significance (** = p<0.01). c) Heatmap depicting changes in expression of selected 22 candidate biomarkers in validation DRESS cohort exposed to culprit drug versus media control. Colour indicates the expression change in logFC. Red: upregulated genes; blue: downregulated genes. d) Heatmap depicting changes in expression of selected 22 candidate biomarkers in validation DRESS cohort versus tolerant patients. Colour indicates the expression change in logFC. Red: upregulated genes, Blue: downregulated genes. e) Box and whisker plot showing cumulative scoring using 22 biomarker genes compared to expected expression alterations based on signature panel. Error bars indicate data range. Horizontal red line indicates threshold score considered positive. (** = p <0.01, Fisher’s exact test). IFN-γ ELIspot = interferon-gamma enzyme-linked immunosorbent spot, SFU = spot forming unit

### An algorithm for analysis of gene expression alterations as a diagnostic approach in antibiotic-DRESS

A point attribution system based on observed changes in each of the transcripts from the 22-gene biomarker panel was developed. Scoring 6 DRESS subjects and 7 tolerant controls showed statistically significant difference (p = 0.0052, Mann-Whitney U test) when scored against all 22 genes (Figure 5e, full scores listed in Supplemental Table S2). By setting a threshold score of 6, this novel scoring system was able to correctly stratify almost all cases (5 DRESS, 6 controls) with high sensitivity and specificity (83.3% and 85.7% respectively, p=0.029, Fisher’s exact test).

### Machine learning identifies optimal panel of biomarkers differentiating antibiotic-DRESS patients from tolerant controls

However, because it was apparent that not all genes contributed equally to the 22-gene scoring matrix that had been developed, we set out to evaluate which gene marker or combination of biomarkers had the highest predictive value for a prospective diagnostic test. Firstly, we took a machine learning approach and trained a random forest algorithm using the validation cohort data (Ranger package, R, alpha = 0.9, trees=500, binary input). The analysis ranked the candidate biomarkers in order of importance for predictive classification (Figure 6a, Supplemental Table S3). For the 10 highest ranked markers, receiver operating characteristics (ROC) analysis showed 100% sensitivity and 100% specificity (AUC = 1). Secondly, we tested a reduced panel of biomarkers identified by their individually significant differential expression between allergics and tolerants: *STAC, GPR183, CD40, CISH, CD4*, and *CCL8* (Figure 6c) in contrast to the other genes in the 22-gene panel (Supplemental Figure 2). By applying our scoring algorithm manually to these 6 genes using a threshold score of 0, we enhanced the diagnostic accuracy as compared to the 22-panel (sensitivity 100%, specificity 85.7%; p = 0.0047, Fisher’s exact test; Figure 6d; Supplemental Table S4).

**Figure 6.**
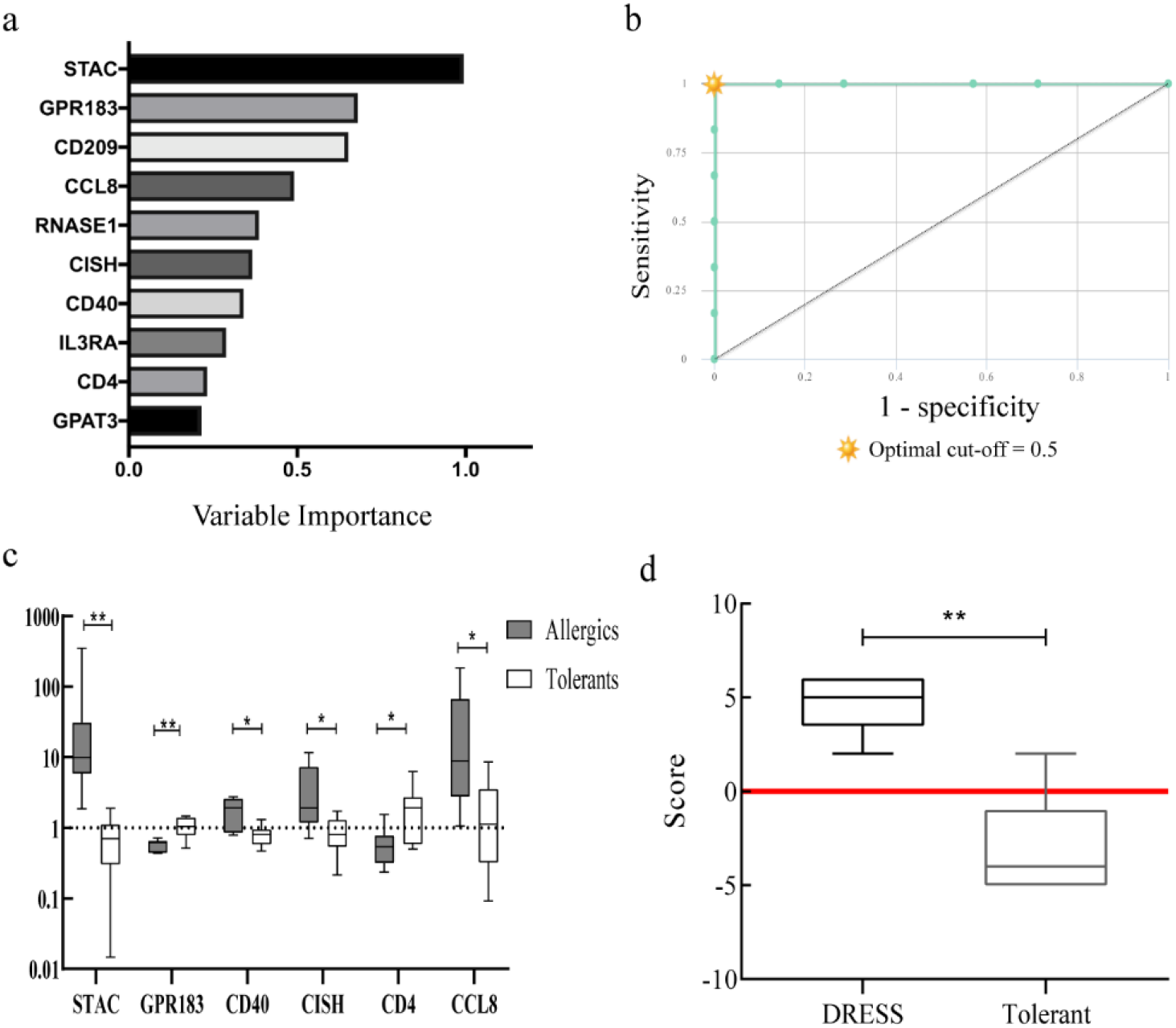
Optimisation of biomarker panel to differentiate DRESS due to antibiotics from tolerant controls. a) Gene importance for the biomarker panel measured by random forest, 10 genes with the highest importance shown (Ranger package, R, alpha = 0.9, trees=500, binary input).b) CombiROC analysis of 10 genes with highest importance (AUC = 1, sensitivity = 100%, specificity = 100%). c) Expression of genes in biomarker panel measured by qPCR in drug allergic patients (grey) and controls tolerant to specified antibiotics (white). Change induced in expression shown for 6 genes reaching statistical difference (* = p <0.05, ** = p < 0.01) in expression change between patient cohorts (2^−ΔΔCT^ versus *YWHAZ* housekeeping gene). Box and whiskers indicate median and data range d) Box and whisker plot of novel scoring system using 6 select biomarker genes to stratify DRESS and control subjects. Error bars indicate data range. Horizontal red line indicates threshold score considered positive. (** = p <0.01, Fisher’s exact test) ROC = receiver operator characteristic, AUC = area under curve, CI = confidence interval.

## Discussion

Criteria for diagnosis of DRESS are clear: cutaneous eruption with hematologic abnormalities and systemic involvement, with the addition of HHV-6 reactivation by Japanese criteria (3, 12). However, the optimal diagnostic work-up to identify a causal drug has remained elusive. Key to the management of DRESS is prompt discontinuation of the culprit drug, as the process can be progressive and even result in catastrophic organ failure (4, 13) and latterly autoimmune sequelae. The determination of drug culpability based only on chronological history of drug ingestion is often unreliable because of heterogeneous presentations and sometimes confusing long-latent periods following the introduction of drugs. In addition to this, definitive challenge testing is inadvisable in DRESS, leaving few alternative options for diagnostic assessment. Particularly challenging are patients taking multiple medications simultaneously. Whilst some groups, including ourselves, have utilised *in vitro* functional T cell assays in an attempt to elucidate the causal drugs (10, 11, 14), multiple issues restrict the widespread availability of such assays. These include the need for specialist resources and expertise, as well as variation in reported sensitivity of tests suggesting a user-dependent variability (14, 15). There is a clear need for new approaches to this issue.

By using a non-hypothesis driven approach to evaluate to identify DRESS activated molecular pathways we sought to maximise the possibility to detect a DRESS-specific signature. Furthermore, such an approach also contributes to better understanding of disease pathogenesis (16, 17). Transcriptomic profiling by RNA sequencing is advantageous as not only does it enable identification of key differentially expressed genes but also has high sensitivity for low abundance transcripts (18, 19). Utilisation of RNAseq in melanoma (20, 21), psoriasis and atopic dermatitis (19) has enabled classification based on phenotype, prognosis, and prediction of intervention outcome. The availability of such technology should therefore be harnessed to further our understanding of cutaneous drug reactions to enable emergent clinical applications.

Here, using a multi-method, unbiased analysis approach, we were able to determine 22 key genes which were differentially regulated in blood cells from allergic individuals after *in vitro* exposure to the culprit drug. Of the 22 transcripts identified, we used a machine learning approach to select 10 and a more traditional differential expression approach to select 6 with the strongest association with DRESS. *GPR183* (G-protein coupled receptor 183; syn. Epstein-Barr virus (EBV)-induced gene 2, *EBI2*) is expressed in lymphocytes where, by binding oxysterols, it creates a chemotactic gradient to direct movement of B-cells, T-cells, dendritic cells and monocytes/macrophages (22, 23). Down-regulation of GPR183 as induced by exposure to the culprit drug in allergics in this study, has been shown to enhance production of type 1 IFNs and inflammatory cytokines by blood dendritic cells (24). Therefore, this may reflect an important pathway for enhanced drug-antigen presentation to CD8+ T cells in DRESS, which may contribute to the organ damage seen in this condition (25). CD4 down-regulation is well established as a consequence of Th2 activation. The down-regulation of CD4 expression in allergics following drug exposure as seen here is interesting because evidence of drug-specific HLA-restriction in DRESS has so far only identified MHC Class I alleles (26). These results therefore support the possibility that drug-specific CD4+ T cells may play an important role in DRESS. Further evidence of the role of CD4 activation is suggested by the enhanced CCL8 expression in allergics. CCL8 has been shown to be central to recruiting IL-5 producing Th2 cells (27), which in turn regulate eosinophilia, thus linking these transcript changes to the hallmarks of DRESS. In addition, *CISH* (cytokine inducible SH2 containing protein), was found to be upregulated by culprit drug exposure in allergics and has been shown to be a marker of allergen-specific Th2 cells (28). Taken together, these data suggest an important role for drug-specific Th2 cells in DRESS and raise the possibility of therapeutic targeting of the Th2 pathway in acute disease. Recent drugs are already licensed for such purposes to treat other Th2 diseases including those targeting IL-4Ra, and anti-IL5. *STAC* (SH3 and cysteine-rich containing protein), a mediator of calcium-dependent inactivation, was also up-regulated in DRESS and whilst it is likely to be important in regulating inflammation (29), the precise role of *STAC1* (as here), remains to be established.

However, for diagnostic approaches, the sensitivity and specificity of the identified signature is key. Using a machine learning approach, we were able to select 10 genes which were demonstrated a sensitivity and specificity of 100%. However, to demonstrate conservative assessment of the utility of these biomarkers in DRESS, we showed that a combined panel of six genes, identified by differential gene expression statistics within the validation cohort allowed robust identification of the causative antibiotic in DRESS with greater accuracy than that of the initial 22 gene algoritm (sensitivity 100%, specificity 85.7%). Furthermore, we showed that these gene expression profiles were not evident in healthy volunteers who tolerated the drugs in question, and also that they were not induced in other inflammatory conditions, which can mimic or precede onset of DRESS. Accordingly, the systems biology approach confirmed that the proposed diagnostic panel is a distinct gene expression signature for DRESS.

Kim et al. recently applied single-cell RNA sequencing (scRNA-seq) to a single case of DRESS, and identified transcriptomal alterations in associated with proliferation, migration, activation and signalling pathways, which then informed therapeutic options (30). Whilst such an approach may be ideal, scRNA-seq applicability to clinical practice is limited by high cost and need for expertise. For diagnostic purposes, we validated that the transcriptomic signature identified here could be reliably interrogated by TaqMan array cards, which would be amenable to utilisation in non-specialist routine pathology laboratories. However, this approach would also be suited to a hub and spoke model for diagnostic testing because the good stability of frozen lysate prior to RNA extraction would enable batched transport and processing if necessary. Whilst downstream processing does require a degree of expertise, centralisation of this would reduce technical variability. It also remains possible that by subjecting greater numbers of cases to molecular profiling, patterns in the gene signature signal could identify hereto-unrecognised DRESS endotypes characterised by a correlation between clinical features and molecular signature (5, 6, 8, 31–34). A wholly *ex vivo* diagnostic test is safe and requires only a minimal amount of blood sampling from patients. Moreover, as the incidence of DRESS is relatively low, between 1:1000 to 1:10,000 drug exposures (35), our preferred approach is to utilise a paired analysis (control vs drug) in diagnostic samples, which mitigates the need for validation of normal ranges for population-wide background correction.

The limitations of this work include the sample size, and the restriction of the allergic cohorts tested to antibiotic induced DRESS. Whilst it remains uncertain whether this transcriptomic signature can be applied to other phenotypes e.g. Stevens-Johnson syndrome, we accepted a high stringency in the clinical phenotype inclusion criteria here because it would minimise sampling error and reduce the size of the cohort required to identify a valid test. Additionally, our tested patients with DRESS were otherwise well at the time of sampling, and therefore, we have no data on the utility of this test in acutely ill patients. However, we did show that the diagnostic panel is not identified in cohorts with other severe infections or other inflammatory skin rashes, suggesting that the gene set measured here is specific to DRESS.

In summary, we show that a carefully selected panel of gene transcripts, which can be measured on a pre-printed array card, offers a useful diagnostic test in antibiotic-associated DRESS with a conservative assessment of 85.7% prediction rate (0.48-0.99 95% CI), and sensitivity of 100% and specificity of 85.7%, but that this could be higher with further refinement and validation. The advantage of this approach is that such gene card testing is familiar to hospital laboratories and therefore this technology is scalable for routine use. Further work is required to determine whether the same panel can be used for other drug hypersensitivity reactions, but the results here demonstrate a proof of concept for the development of gene signature panels for diagnosis of T cell mediated disease.

## Methods

### Patients and controls

Patients diagnosed with delayed hypersensitivity reaction (DHR) on clinical grounds by consultant dermatologists experienced in the recognition of these reactions were recruited to the study through the Department of Dermatology, University Hospital Southampton NHS Foundation Trust. Eleven DRESS patients with symptoms corresponding to diagnostic criteria for DRESS were identified as confirmed by RegiSCAR score ≥3 (4). Only subjects with no active infections or malignancies and without history of immunosuppression, including due to medications, were included. 7 comparative tolerant controls (treated with a relevant antibiotic, but without DHR symptoms) were also tested. Patients were divided into a ‘discovery’ cohort (5 patients with DRESS caused by antibiotics: cefoxitin, dapsone, teicoplanin, or vancomycin) and a ‘validation’ cohort (6 patients with DRESS) (Table 1). All testing was undertaken on fresh (not frozen) samples isolated from anticoagulated peripheral blood. The tests were undertaken on average (mean) 370.7 days from rash onset (median: 124 days, IQR 71-347).

### Lymphocyte Proliferation and ELISpot test

Lymphocyte proliferation test and IFN-γ enzyme-linked immunosorbent spot assay (ELISpot) are routinely performed in our laboratory as we previously described (10, 11). Briefly, peripheral blood mononuclear cells (PBMCs) were cultured in the presence of the relevant drug at four concentrations overnight at 37°C, 5% CO_2_on pre-coated ELISpot plates (Millipore, UK; Mabtech, Sweden) and for 5 days (lymphocyte proliferation assay, LPA) respectively, in triplicate. Internal validation of the assays was achieved with negative (medium) and positive (phytohemagglutinin, PHA) controls. For ELISpot testing, after overnight culture, plates were washed and developed as per manufacturer’s instructions. Spot-forming units per million cells were enumerated using an automated ELISpot reader (Autoimmun Diagnostika GmbH, Strassberg, Germany). Positive responses were recorded as those responses greater than the mean of all the background samples plus 2x the standard deviation (SD) of the background. For LPA testing, on day 5, 1 μCi/ml of 3H-thymidine (Amersham, UK) was added, and the cells harvested 6 hours later for scintillation counting. The stimulation index (SI) was calculated as the fold difference between counts per minute recorded in wells stimulated by drug over the negative control. SI greater than 2 was considered a positive result (Table 1).

### RNA isolation and purification

PBMCs (7.5×10^5^ cells per well, in duplicates) were incubated for 24 hours with medium (control) or culprit drug before RNA harvesting for transcriptomic analysis. Following 24-hour stimulation with drug or control medium, PBMCs were harvested, washed, and suspended in RLT lysing buffer (Qiagen, UK) before storage at −20 °C. Each sample was thawed immediately before RNA isolation and whole transcriptome RNA-sequencing. RNA extraction and purification were performed according to manufacturer’s protocol (RNeasy Plus Mini Kit, Qiagen, UK). DNA contamination in the collected RNA was eliminated by use of gDNA Eliminator spin column. RNA quantity and quality checking were performed using the NanoDrop™ 1000 spectrophotometer (Thermo Fisher Scientific, Waltham, MA, USA) and Agilent 2100 Bioanalyzer using an RNA 6000 Nano LabChip kit (Agilent Technologies, Cork, Ireland). All samples displayed a 260/280 ratio >1.8 and RNA integrity numbers (RIN) > 7.7. Purified RNA samples were stored at −80 °C until use.

### mRNA-Seq library construction and sequencing

Total RNA samples were subjected to indexed cDNA library construction, using the Illumina TruSeq poly(A) + RNA-Seq library construction, according to the manufacturer’s instructions. For sequencing, all samples were pooled in a single pool and sequenced on 3 lanes, yielding 75-bp paired-end reads, using an Illumina HiSeq 4000 platform (an outsourced service at the Oxford Genomics Centre).

### Bioinformatics analysis

Quality-controlled reads were aligned to the reference genome GRCh37.EBVB95-8wt.ERCC using the HISAT aligner. Alignments were counted for each gene using the featureCounts package (36). Aligned reads were further analysed in R using the Bioconductor suite of packages. Filtered trimmed mean of M values (TMM) normalised counts per million (cpm) (EdgeR, filtering out genes less than two gene counts in at least half of the samples) were used for downstream analyses (37). Determination of differentially expressed genes (DEG) was performed using EdgeR with a nested paired design (37). The expected false discovery rate (FDR) was estimated using the Benjamini-and-Hochberg method. An FDR adjusted p ≤0.05 was considered significant.

RNA-seq data were deposited in accordance with MIAME guidelines, in Gene Expression Omnibus (GEO) under accession number GSE160369.

### Quantitative Reverse Transcription-PCR (qRT-PCR)

The expression of chosen genes was validated with quantitative PCR using the TaqMan gene expression assays for target genes: *YWHAZ* (Hs01122445_g1), *STAC* (Hs00182385_m1), *CISH* (Hs00367082_g1), *FN1* (Hs01549976_m1) and *CD4* (Hs01058407_m1) (Applied Biosystems, Life Technologies, UK) in PBMCs isolated from whole blood. RNA extraction (RNeasy Plus Mini Kit, Qiagen) and cDNA reverse transcription, including RT-negative control, (High-Capacity cDNA Reverse Transcription Kit, Applied Biosystems; ThermoFisher Scientific UK) were carried out according to the manufacturer’s protocol. qPCR was performed in 384-well plate assay, using Applied Biosystems 7900HT Fast Real-Time PCR System. Gene expression levels were normalised to housekeeping gene expression (*YWHAZ*).

### TaqMan array card

Customised RT-PCR cards from Applied Biosystems (http://www.appliedbiosystems.com) were used in the quantitative analysis of the 22 selected candidate genes. Eight samples with two technical duplicates were tested per card. The 384-well microfluidic card was preloaded with our chosen genes. Each cDNA sample was added to an equal volume of mastermix (TaqMan, Applied Biosystems) and then loaded onto the array card. PCR amplification was performed using a 7900HT Fast Real-time PCR System (Applied Biosystems) following the protocol described by the manufacturer. The threshold cycle (CT) was automatically given by the SDS2.2® software package (Applied Biosystems). Relative expression of each gene was normalized to *YWHAZ* as the sole housekeeping gene, and log2-transformed for analysis (RQ = 2^−ΔΔCt^). All data were generated in duplicate for each gene expression per sample.

### Evaluation of diagnostic performances

Ranking of detected genes for selection of candidate biomarker genes was done using absolute log fold change (FC) cut off (logFC≥|1.5|) calculated using generalised linear model in EdgeR, combined with minimum expression levels for all donors (minimum cpm≥4, maximum cpm≥100). Random forest analysis was performed using package Ranger in R (importance measure = impurity, number of trees = 500, alpha = 0.9). Combinatorial panel analysis with top 10 candidate genes identified on random forest algorithm were performed using CombiRoc webtool (38). Receiver-operating characteristic (ROC) curves were calculated in order to assess the diagnostic power of the gene combination by the area under the curve (AUC) of the ROC curve. Potential biomarkers were considered valuable if sensitivity and specificity were >85%, as well as AUC ≥0.8.

### Comparison with systemic inflammatory conditions

Datasets for 4 systemic conditions: influenza, sepsis, systemic lupus erythematosus and dermatomyositis were downloaded from publically available genomic data repositories (GSE114588, GSE60424, GSE112087, GSE125977). FASTQ files for GSE114588 and GSE60424 were aligned using Kallisto (39) against the GRCh38 human reference genome followed by differential analysis using Sleuth (40). Disease describing gene expression signatures were generated by comparing TMM normalised gene expression levels between experimental and control group using EdgeR package (37) (FC ≥log2 and adjusted p value <0.05). Raw RNA-seq data for GSE112087 was quantified to gene-level counts using the ARCHS4 pipeline (41) with similar thresholds as the other datasets. Published values (FC ≥log2) relating to dermatomyositis subjects from GSE125977 were extracted for comparative analysis. Enrichment analyses performed to published gene sets associated with these four inflammatory conditions (influenza 2, sepsis 5, systemic lupus erythematosus 5, dermatomyositis 1) did not show significant overlap (enrichment scores: 0.27-0.55, FDR<0.05).

### Functional enrichment analysis

Gene set enrichment analysis (GSEA) (42, 43) was performed for complete DRESS dataset (11747 transcripts, average were calculated for transcripts associated with the same genes (3 genes)) using curated gene signatures of 4 inflammatory diseases above downloaded from MSigDB (Molecular Signatures Database v7.1) (**Supplemental Table S5**). Largest collections relating to dermatomyositis from DisGeNET platform (v7.0) (44, 45) were used in view of no available curated gene sets for this disease on other MSigDB platform (42, 46). Similarities were examined at cut-off of FDR-adjusted p-value <0.05 and Enrichment Scores.

### Scoring classification

Mean values for each biomarker gene was calculated from RT-qPCR data from 6 DRESS subjects tested using the array card and compared against logFC RNAseq data to determine up- and down-regulated genes in the signature panel. For every transcript expression which matched this expected change, 1 point was added whilst 1 point was subtracted if direction of change was opposite to that of the identified signature. Log2 2^−ΔΔCt^ values for each subject (6 DRESS, 7 tolerant controls) were used in this scoring. No points were added or subtracted if values fell between −0.25 and 0.25.

### Statistics

Statistical analyses were performed using Prism 8.1 (GraphPad Software) and methods embedded in bioinformatics pipelines (Generalised Linear Model, EdgeR, Benjamini-and-Hochberg FDR-corrected p-value test). Mann-Whitney U test was used for comparison between non-matched non-parametric samples and Fisher’s exact test for contingency table analysis. Correlations between RNA-seq and qPCR results were performed using Pearson test and linear regression analysis. Data were considered significant at p<0.05.

### Study approval

Ethical approval for this project was obtained from the UK Research Ethics Service (RES) of the national Health Research Authority (17/NE/0346) for subject recruitment at University Hospital Southampton NHS Trust. Written informed consent from all participants was obtained prior to inclusion in the study.

## Data Availability

https://www.ncbi.nlm.nih.gov/geo/query/acc.cgi?acc=GSE160369

## Author contributions

YXT and WYH performed experiments, analysed data and drafted the manuscript. AFV analysed data and provided technical support. CG performed experiments. JW analysed data. PSF contributed to study design and oversaw writing of the manuscript. MEP and MAJ designed the research study, analysed data and oversaw writing of the manuscript.

## Acknowledgements

This work was supported by the British Skin Foundation [grant number BSF8021]. MEP is funded by the Wellcome Trust [grant number 109377/Z/15/Z]. The authors acknowledge the use of the IRIDIS High Performance Computing Facility, and associated support services at the University of Southampton, in the completion of this work as well as assistance from Christopher Woelk PhD, and Tilman Sanchez-Elsner PhD. We thank the Oxford Genomics Centre at the Wellcome Centre for Human Genetics (funded by Wellcome Trust grant reference 203141/Z/16/Z) for the generation and initial processing of the sequencing data.

