## Supplemental data for "Molecular diagnosis of causality in T cell mediated severe cutaneous adverse drug reactions"

**Conflict of interest:** The authors have declared that no conflict of interest exists

**Keywords:**

Drug reaction with eosinophilia and systemic symptoms (DRESS); Drug allergy; Immunology; Biomarker; Diagnostics

[1] Clinical Experimental Sciences, Faculty of Medicine, University of Southampton, Southampton SO16 6YD, United Kingdom

[2] Department of Dermatology, Southampton General Hospital, University Hospitals Southampton NHS Foundation Trust

**Supplemental Figure 1**

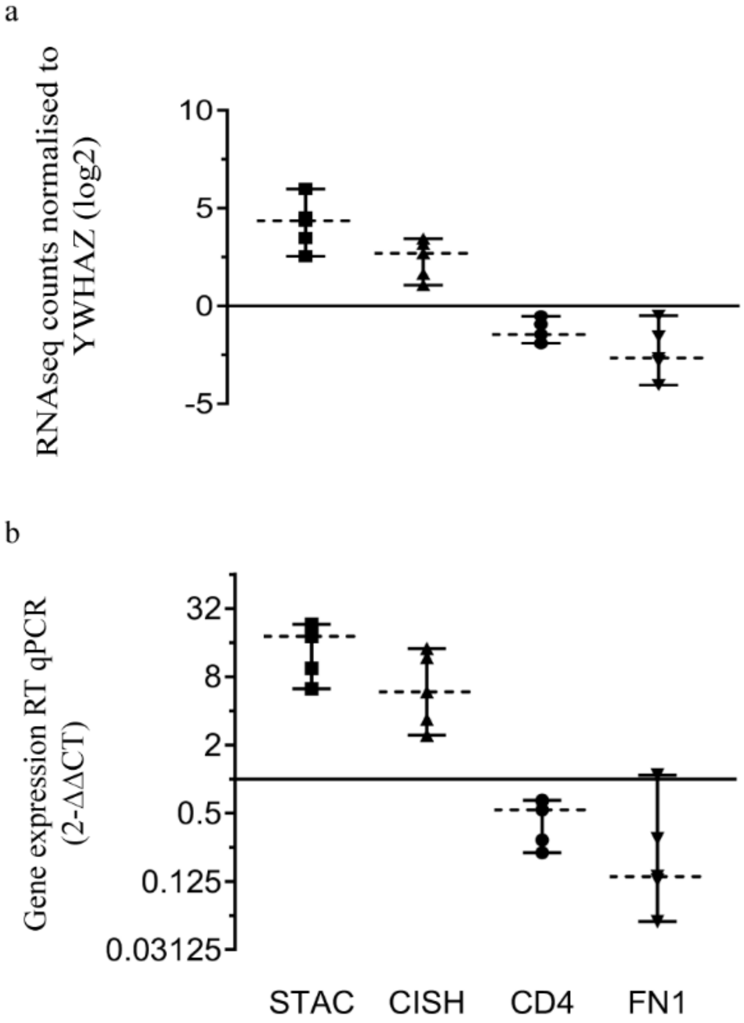

**Supplemental Figure 1.** Validation of top DEGs identified by whole transcriptome analysis. Changes in gene expression detected by RNA-sequencing were reproduced using RT-qPCR for the top 4 gene transcripts. a) Gene expression measured using RNA-sequencing for *STAC*, *CISH*, *CD4* and *FN1*. Log2 of TMM normalised counts calibrated to *YWHAZ* b) Gene expression measured using RT-qPCR for *STAC*, *CISH*, *CD4* and *FN1* $2^{-\Delta\Delta CT}$, *YWHAZ* as housekeeping gene, in duplicate. Each data point represents gene expression in a single patient. Horizontal dotted line shows group median, error bars depict data range.

**Table S1:** Genes in biomarker panel

| Gene name | Gene symbol | Assay ID |
|---|---|---|
| Housekeeping genes | | |
| Tyrosine 3-monooxygenase/tryptophan 5-monooxygenase activation protein zeta | YWHAZ | Hs01122445_g1 |
| Glyceraldehyde-3-Phosphate Dehydrogenase | GAPDH | Hs99999905_m1 |
| RNA-seq selected DEGs | | |
| SH3 and cysteine rich domain | STAC | Hs00182385_m1 |
| Inhibin beta A subunit | INHBA | Hs01081598_m1 |
| C-C motif chemokine ligand 8 | CCL8 | Hs04187715_m1 |
| Ribonuclease A family member 1, pancreatic | RNASE1 | Hs01850125_s1 |
| Cytokine inducible SH2 containing protein | CISH | Hs00367082_g1 |
| Cathepsin L | CTSL | Hs00964650_m1 |
| Mannose receptor, C type 1 | MRC1 | Hs00267207_m1 |
| Heparin binding EGF like growth factor | HBEGF | Hs00181813_m1 |
| Fibronectin 1 | FN1 | Hs01549976_m1 |
| Glycerol-3-phosphate acyltransferase 3 | GPAT3 | Hs00262010_m1 |
| Interleukin 3 receptor subunit alpha | IL3RA | Hs00608141_m1 |
| CD274 molecule | CD274 | Hs00204257_m1 |
| CD40 molecule | CD40 | Hs01002915_g1 |
| CD276 molecule | CD276 | Hs00987207_m1 |
| Integrin subunit alpha M | ITGAM | Hs00167304_m1 |
| CD209 molecule | CD209 | Hs01588349_m1 |
| Carboxypeptidase, vitellogenic like | CPVL | Hs01073862_m1 |
| CD4 molecule | CD4 | Hs01058407_m1 |
| G protein-coupled receptor 183 | GPR183 | Hs00270639_s1 |
| V-Set Immunoregulatory Receptor | VSIR | Hs00735289_m1 |
| Tumor necrosis factor | TNF | Hs00174128_m1 |
| Interferon gamma | IFNG | Hs00989291_m1 |

**Table S2:** Scoring using 22 biomarker genes in identification of DRESS subjects

| **Biomarker** | DRESS 1 | DRESS 2 | DRESS 3 | DRESS 4 | DRESS 5 | DRESS 6 | Tolerant 1 | Tolerant 2 | Tolerant 3 | Tolerant 4 | Tolerant 5 | Tolerant 6 | Tolerant 7 |
|---|---|---|---|---|---|---|---|---|---|---|---|---|---|
| CTSL | 1 | 1 | 1 | 1 | 1 | 1 | 0 | 1 | -1 | -1 | -1 | -1 | 1 |
| IL3RA | 1 | 1 | 1 | 1 | 1 | -1 | -1 | 0 | 0 | -1 | -1 | -1 | 1 |
| CISH | 1 | 1 | -1 | 1 | 1 | 1 | -1 | 1 | 0 | -1 | -1 | -1 | -1 |
| TNF | 0 | 1 | 1 | 1 | 1 | -1 | 0 | 1 | -1 | 0 | 0 | -1 | 1 |
| ITGAM | 1 | 1 | -1 | 0 | 1 | -1 | -1 | 1 | 1 | 1 | 1 | -1 | 1 |
| HBEGF | 0 | 1 | 1 | 1 | 1 | -1 | 1 | 1 | 0 | 1 | 1 | -1 | 0 |
| CD276 | 1 | 0 | 1 | 1 | 1 | -1 | -1 | 0 | -1 | 1 | 1 | 0 | 0 |
| INHBA | 1 | 1 | 1 | 1 | 1 | -1 | 1 | 1 | 0 | -1 | -1 | -1 | 0 |
| GPAT3 | 1 | 0 | 1 | 1 | 1 | 0 | 0 | 1 | 1 | 1 | 1 | 1 | 1 |
| CCL8 | 1 | 1 | 1 | 1 | 1 | 1 | 1 | 1 | -1 | -1 | -1 | 0 | 1 |
| CD274 | 1 | 1 | -1 | 1 | 1 | 1 | 1 | 1 | 0 | -1 | -1 | -1 | 1 |
| CD209 | -1 | 1 | 0 | -1 | 1 | 1 | 1 | -1 | -1 | 0 | 0 | 1 | -1 |
| CD40 | 0 | 1 | -1 | 1 | 1 | 1 | -1 | 0 | -1 | -1 | -1 | 0 | 1 |
| MRC1 | -1 | 1 | -1 | -1 | 1 | 1 | 1 | 1 | 1 | -1 | -1 | -1 | -1 |
| STAC | 0 | 1 | 1 | 1 | 1 | 1 | -1 | -1 | 0 | 0 | 0 | -1 | -1 |
| IFNG | 1 | 1 | -1 | 1 | 1 | -1 | -1 | 1 | -1 | -1 | -1 | 1 | 1 |
| GPR183 | 1 | 1 | 1 | 1 | 1 | 1 | -1 | -1 | -1 | -1 | -1 | -1 | 1 |
| RNASE1 | 1 | 1 | 1 | 1 | 1 | 1 | -1 | -1 | 1 | -1 | -1 | 1 | 1 |
| FN1 | 1 | 1 | 1 | 1 | 1 | 1 | -1 | -1 | 1 | -1 | -1 | -1 | 1 |
| CPVL | 1 | -1 | -1 | 1 | 1 | -1 | -1 | 1 | -1 | -1 | -1 | 1 | -1 |
| CD4 | 1 | -1 | 1 | 1 | 1 | 1 | -1 | -1 | -1 | -1 | -1 | 1 | 1 |
| VSIR | 1 | 1 | 1 | 1 | -1 | 1 | -1 | -1 | 1 | -1 | -1 | -1 | 1 |
| **Score** | **14** | **16** | **7** | **17** | **20** | **5** | **-7** | **5** | **-4** | **-11** | **-11** | **-7** | **9** |

**Supplemental Figure 2**

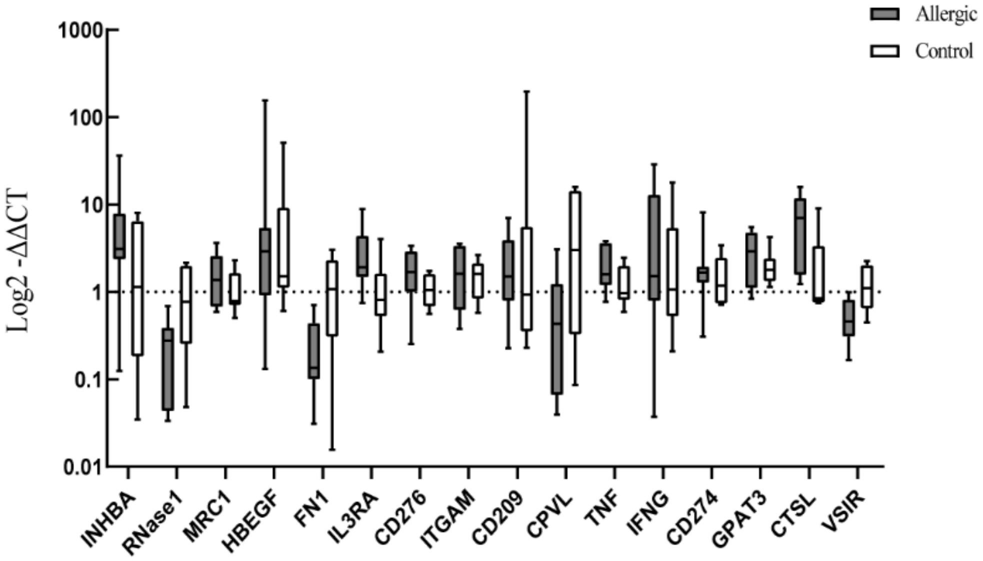

**Supplementary Figure 2.** Expression of remaining genes from gene panel in validation cohort and in tolerant subjects. Expression of genes in biomarker panel measured by qPCR in drug allergic patients (grey) and controls tolerant to specified antibiotics (white). Change induced in expression shown for genes not reaching statistical difference of p = <0.05 in expression change between patient cohorts ($2^{-\Delta\Delta CT}$ versus *YWHAZ* housekeeping gene). Box and whiskers indicate median and data range.

**Table S3:** Biomarker genes ranked according to random forest algorithm values.

| | variable importance |
|---|---|
| STAC | 0.994811833 |
| GPR183 | 0.680540981 |
| CD209 | 0.651630914 |
| CCL8 | 0.49121443 |
| RNASE1 | 0.386492208 |
| CISH | 0.36682482 |
| CD40 | 0.341145022 |
| IL3RA | 0.289051948 |
| CD4 | 0.233019336 |
| GPAT3 | 0.215453824 |
| CD276 | 0.185681962 |
| CTSL | 0.172102742 |
| IFNG | 0.171135786 |
| FN1 | 0.167650938 |
| MRC1 | 0.161882251 |
| ITGAM | 0.161407648 |
| C10orf54 | 0.157426018 |
| CD274 | 0.12821746 |
| TNF | 0.113121756 |
| HBEGF | 0.110421068 |
| INHBA | 0.096309812 |

**Table S4:** Scoring using 6 selected biomarker genes in identification of DRESS subjects

| **Biomarker** | DRESS 1 | DRESS 2 | DRESS 3 | DRESS 4 | DRESS 5 | DRESS 6 | Tolerant 1 | Tolerant 2 | Tolerant 3 | Tolerant 4 | Tolerant 5 | Tolerant 6 | Tolerant 7 |
|---|---|---|---|---|---|---|---|---|---|---|---|---|---|
| CISH | 1 | 1 | -1 | 1 | 1 | 1 | -1 | 1 | 0 | -1 | -1 | -1 | -1 |
| CCL8 | 1 | 1 | 1 | 1 | 1 | 1 | 1 | 1 | -1 | -1 | -1 | 0 | 1 |
| CD40 | 0 | 1 | -1 | 1 | 1 | 1 | -1 | 0 | -1 | -1 | -1 | 0 | 1 |
| STAC | 0 | 1 | 1 | 1 | 1 | 1 | -1 | -1 | 0 | 0 | 0 | -1 | -1 |
| GPR183 | 1 | 1 | 1 | 1 | 1 | 1 | -1 | -1 | -1 | -1 | -1 | -1 | 1 |
| CD4 | 1 | -1 | 1 | 1 | 1 | 1 | -1 | -1 | -1 | -1 | -1 | 1 | 1 |
| **Score** | **4** | **4** | **2** | **6** | **6** | **6** | **-4** | **-1** | **-4** | **-5** | **-5** | **-2** | **2** |

**Table S5:** Inflammatory diseases gene expression signature overlap with biomarker panel analysis

| Disease | Curated gene set (GEO dataset) | Gene expression in curated gene set | Number of genes | Normalised enrichment score | FDR q-value | Overlap genes with DRESS biomarker |
|---|---|---|---|---|---|---|
| Influenza | GSE6269: healthy vs influenza PBMC | Upregulated | 160 | 1.458515 | 0.006908463 | |
| Sepsis | GSE9960: healthy vs sepsis PBMC | Upregulated | 192 | 2.433338 | 0.0 | STAC |
| | GSE9960 healthy vs gram positive sepsis PBMC | Upregulated | 198 | 2.1894507 | 0.0 | INHBA, CD274 |
| | GSE9960 healthy vs gram negative sepsis PBMC | Downregulated | 198 | 2.4648156 | 0.0 | INHBA |
| | GSE9960 healthy vs gram negative and positive sepsis PBMC | Downregulated | 196 | 1.4939677 | 0.004305705 | CCL8 |
| Systemic lupus erythematosus | GSE10325 myeloid vs lupus | Upregulated | 190 | 2.4966972 | 0.0 | CCL8 |
| | GSE10325 CD4 T cell vs lupus CD4 T cells | Upregulated | 157 | 2.166506 | 0.0 | |
| | Bennett systemic lupus erythematosus M12175 | Upregulated | 27 | 1.3876796 | 0.04183008 | |
| | GSE30153 lupus vs healthy donor B cells | Upregulated | 146 | 1.1878923 | 0.1454361 | |
| | GSE10325 myeloid vs lupus myeloid | Downregulated | 70 | 1.2539403 | 0.10766052 | |
| Dermatomyositis | Disgenet dermatomyositis CUI: C0011633 | Upregulated | 149 | 1.5843996 | 0.0034364262 | TNF, CD274, CTSL, MRC1 |

DRESS = drug reaction with eosinophilia and systemic symptoms; FDR = false discovery rate; GEO = gene expression omnibus; PBMC = peripheral blood mononuclear cells; vs = versus
